# Feasibility of Controlling COVID-19 Outbreaks in the UK by Rolling Interventions

**DOI:** 10.1101/2020.04.05.20054429

**Authors:** Po Yang, Jun Qi, Shuhao Zhang, Xulong wang, Gaoshan Bi, Yun Yang, Bin Sheng, Xuxin Mao

**Affiliations:** Department of Computer Science, The University of Sheffield, Sheffield, United Kingdom; Department of Engineering Science, University of Oxford, Oxford, United Kingdom; Oxford Suzhou Centre for Advanced Research, Suzhou, China; School of Software, Yunnan University, Yunnan, China; Department of Computer Science and Engineering, Shanghai JiaoTong University, Shanghai, China; National Institute of Economic and Social Research, London, United Kingdom; European Institute, London School of Economics and Political Science, London, United Kingdom

## Abstract

**Background:** Recent outbreak of a novel coronavirus disease 2019 (COVID-19) has led a rapid global spread around the world. For controlling COVID-19 outbreaks, many countries have implemented two non-pharmaceutical interventions: suppression like immediate lock-downs in cities at epicentre of outbreak; or mitigation that slows down but not stopping epidemic for reducing peak healthcare demand. Both interventions have apparent pros and cons; the effectiveness of any one intervention in isolation is limited. We aimed to conduct a feasibility study for robustly estimating the number and distribution of infections, growth of deaths, peaks and lengths of COVID-19 breakouts by taking multiple interventions in London and the UK, accounting for reduction of healthcare demand.

**Methods:** We developed a model to attempt to infer the impact of mitigation, suppression and multiple rolling interventions for controlling COVID-19 outbreaks in London and the UK. Our model assumed that each intervention has equivalent effect on the reproduction number R across countries and over time; where its intensity was presented by average-number contacts with susceptible individuals as infectious individuals; early immediate intensive intervention led to increased health need and social anxiety. We considered two important features: direct link between Exposed and Recovered population, and practical healthcare demand by separation of infections into mild and critical cases. Our model was fitted and calibrated with data on cases of COVID-19 in Wuhan to estimate how suppression intervention impacted on the number and distribution of infections, growth of deaths over time during January 2020, and April 2020. We combined the calibrated model with data on the cases of COVID-19 in London and non-London regions in the UK during February 2020 and March 2020 to estimate the number and distribution of infections, growth of deaths, and healthcare demand by using multiple interventions.

**Findings:** We estimated given that multiple interventions with an intensity range from 3 to 15, one optimal strategy was to take suppression with intensity 3 in London from 23^rd^ March for 100 days, and 3 weeks rolling intervention with intensity between 3 and 5 in non-London regions. In this scenario, the total infections and deaths in the UK were limited to 2.43 million and 33.8 thousand; the peak time of healthcare demand was due to the 65^th^ day (April 11^th^), where it needs hospital beds for 25.3 thousand severe and critical cases. If we took a simultaneous 3 weeks rolling intervention with intensity between 3 and 5 in all regions of the UK, the total infections and deaths increased slightly to 2.69 million and 37 thousand; the peak time of healthcare kept the same at the 65^th^ day, where it needs equivalent hospital beds for severe and critical cases of 25.3 thousand. But if we released high band of rolling intervention intensity to 6 or 8 and simultaneously implemented them in all regions of the UK, the COVID-19 outbreak would not end in 1 year and distribute a multi-modal mode, where the total infections and deaths in the UK possibly reached to 16.2 million and 257 thousand.

**Interpretation:** Our results show that taking rolling intervention is probably an optimal strategy to effectively and efficiently control COVID-19 outbreaks in the UK. As large difference of population density and social distancing between London and non-London regions in the UK, it is more appropriate to implement consistent suppression in London for 100 days and rolling intervention in other regions. This strategy would potentially reduce the overall infections and deaths, and delay and reduce peak healthcare demand.

**Research in context:** *Evidence before this study:* Suppression and mitigation are two common interventions for controlling infectious disease outbreaks. Previous works show rapid suppression is able to immediately reduce infections to low levels by eliminating human-to-human transmission, but needs consistent maintenance; mitigation does not interrupt transmission completely and tolerates some increase of infections, but minimises health and economic impacts of viral spread.^3^ While current planning in many countries is focused on implementing either suppression or mitigation, it is not clear how and when to take which level of interventions for control COVID-19 breakouts to certain country in light of balancing its healthcare demands and economic impacts.

*Added value of this study:* We used a mathematical model to access the feasibility of multiple intervention to control COVID-19 outbreaks in the UK. Our model distinguished self-recovered populations, infection with mild and critical cases for estimating healthcare demand. It combined available evidence from available data source in Wuhan. We estimated how suppression, mitigation and multiple rolling interventions impact on controlling outbreaks in London and non-London regions of the UK. We provided an evidence verification point that implementing suppression in London and rolling intervention with high intensity in non-London regions is probably an optimal strategy to control COVID-19 breakouts in the UK with minimised deaths and economic impacts.

*Implications of all the available evidence:* The effectiveness and impact of suppression and mitigation to control outbreaks of COVID-19 depends on intervention intensity and duration, which remain unclear at the present time. Using the current best understanding of this model, implementing consistent suppression in London for 100 days and 3 weeks rolling intervention with intensity between 3 and 5 in other regions potentially limit the total deaths in the UK to 33.8 thousand. Future research on how to quantify and measure intervention activities could improve precision on control estimates.

## Introduction

As of April 1st, 2020, the ongoing global epidemic outbreak of coronavirus disease 2019 (COVID-19) has spread to at least 146 countries and territories on 6 continents, resulted in 896 thousands confirmed case and over 45 thousands deaths.^1^ In the UK, COVID-19 infections and deaths reached 29478 and 2352, with a mortality ratio nearly 7.9%.^1^ For effectively controlling COVID19-breaks, most countries have implemented two non-pharmaceutical interventions: suppression strategy like immediate lockdowns in some cities at epicentre of outbreak; or mitigation that slows down but not stopping epidemic for reducing peak healthcare demand.^2.3.4^

However, both above interventions have apparent pros and cons; the effectiveness of any one intervention in isolation is limited.^4.^ Taking an example of controlling the COVID-19 epidemic in Wuhan, suppression strategy with extremely high intensity (the highest state of emergency) were token by China government from 23^nd^ January 2020 for 50 days, resulting prevention of over 700 thousand national infectious case.^5^ However, China’s first quarter gross domestic product is estimated to a year-on-year contraction to 9 percent.^6^ In most scenarios, it is difficult to conduct an optimal intervention that minimises both growing infections and economic loss in ongoing COVID-19 breakouts.

The effectiveness of intervention strategies is accessed by decline of daily reproduction parameter R_t_, that used to measure a transmission potential of a disease. The R_t_ of COVID-19 is estimated to be 2.5 -3.2. ^7,8.9.10^ Its implementation hinges on two parameters: intervention intensity presented by average-number contacts per person, and intervention duration counted by weeks.^11^ The practical impacts of applying intervention strategies to certain country are varied in light of many factors including population density, human mobility, health resources, culture issues, etc. It is crucial but hard to know how and when to take which level of interventions tailored to the specific situation in each country.

Targeting at this problem, we aimed to conduct a feasibility study that explored a range of epidemiological scenarios by taking different intervention strategies on current information about COVID-19 outbreaks in the UK. We assessed the effectiveness of multiple interventions to control outbreaks using a mathematical transmission model accounting for available and required healthcare resources by distinguishing self-recovered populations, infection with mild and critical cases. By varying the intensity, timing point, period and combinations of multiple interventions, we show how viable it is for the UK to minimise the total number of infections and deaths, delay and reduce peak of healthcare demand.

## Methods

### Mode structure

We implemented a modified SEIR model to account for a dynamic Susceptible [S], Exposed [E] (infected but asymptomatic), Infectious [I] (infected and symptomatic) and Recovered [R] or Dead [D] population’s state. For estimating healthcare needs, we categorised infectious group into two sub-cases: Mild [M] and Critical [C]; where Mild cases did not require hospital beds; Critical cases need hospital beds but possibly cannot get it due to shortage of health sources. Conceptually, the modified modal is shown in Fig.2.

**Figure 1:**
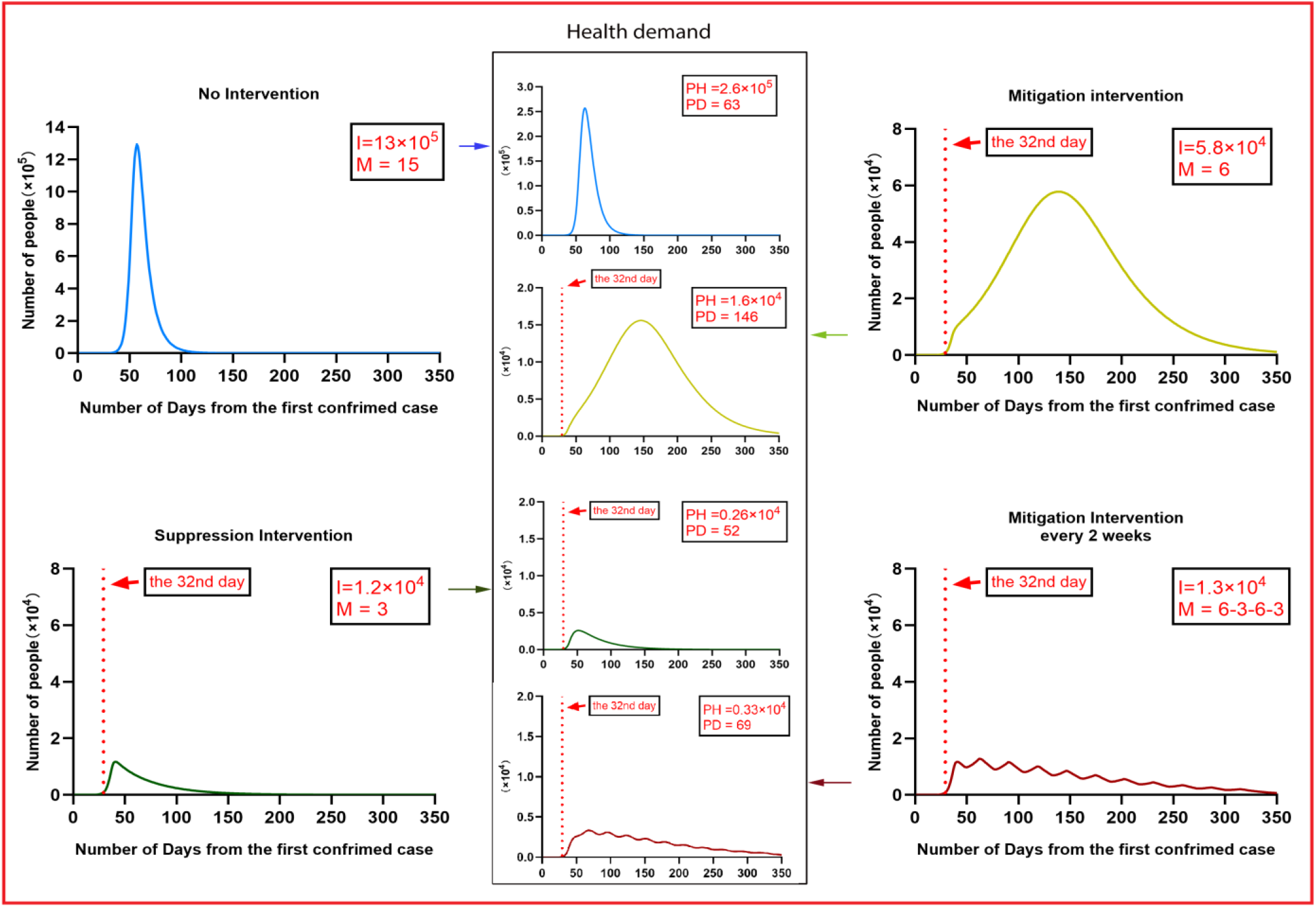
Illustration of controlling Wuhan COVID-19 outbreaks by taking different intervention strategies with parameters. (City populations: 1.4 million; daily contacts per person: No Interventions (M = 15), Suppression Intervention (M = 3), Mitigation Intervention (M = 6), and Hybrid intervention (6-3-6-3) every two weeks; I: Peak daily infections; PH: Peak value of health demand, PD: Peak date of health demand).

**Figure 2:**
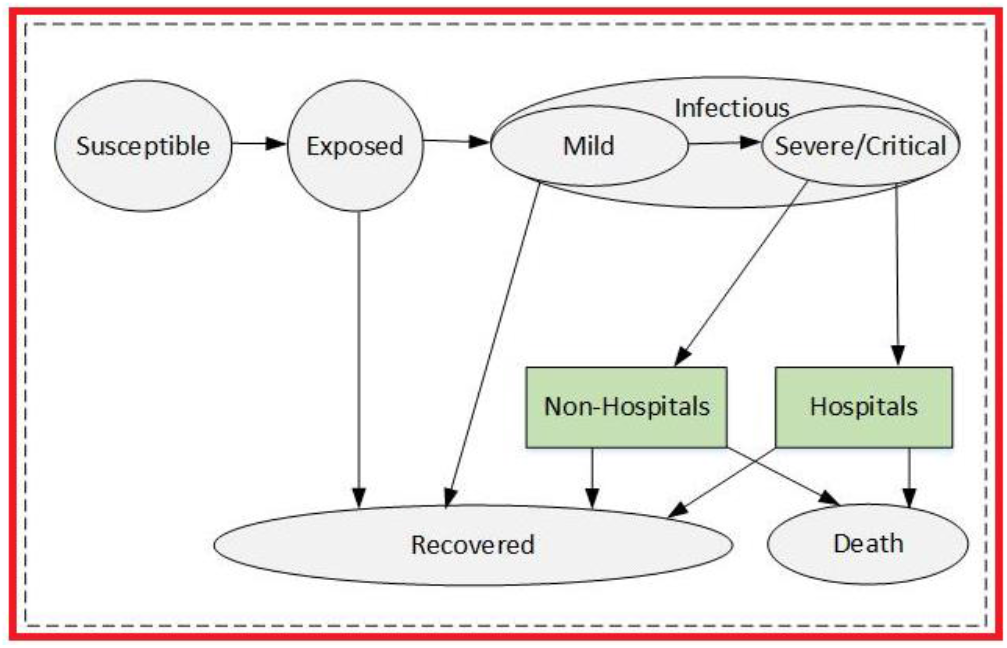
Extended SEMCR model structure: The population is divided into the following six classes: susceptible, exposed (and not yet symptomatic), infectious (symptomatic), mild (mild or moderate symptom), critical (severe symptom), death and recovered (i.e, isolated, recovered, or otherwise non-infectious).

The model accounted for delays in symptom onset and reporting by including compartments to reflect transitions between reporting states and disease states. Here, this modal assumed that S is initial susceptible population of certain region; and incorporated an initial intervention of surveillance and isolation of cases in contain phase by a parameter β.^14.15^ If effectiveness of intervention in contain phase was not sufficiently strong, susceptible individuals may contract disease with a given rate when in contact with a portion of exposed population E. After an incubation period *α*_1_, the exposed individuals became the infectious population I at a ratio 1/*α*_1_. The incubation period was assumed to be 5.8 days.^8^ Once exposed to infection, infectious population started from Mild cases M to Critical cases C at a ratio *a*, Critical cases led to deaths at a ratio *d*; other infectious population finally recovered. We assumed that COVID-19 can be initially detected in 2 days prior to symptom onset and persist for 7 days in moderate cases and 14 days to severe cases.^19^

Notably, two important features in our model differ with other SIR or SEIR models.^12.13^ The first one was that we built two direct relationships between Exposed and Recovered population, Infections with mild symptoms and Recovered population. It was based on an observation of COVID-19 breakouts in Wuhan that a large portion (like 42.5% in Wuhan) of self-recovered population were asymptomatic or mild symptomatic.^14^ They did not go to hospital for official COVID-19 tests but actually were infected. Without considering this issue, the estimation of total infections were greatly underestimated.^13^ In order to measure portion of self-recovery population, we assumed that exposed individuals at home recovered in 3.5 days; mild case at home recovered in 7 days.^19^

The second feature was to consider shortage of health sources (hospital beds) in the early breakouts of COVID-19 might lead to more deaths, because some severe or critical cases cannot be accommodated in time and led to death at home (non-hospital). For instance, in Wuhan, taking an immediate suppression intervention on 23^rd^ Jan 2020 increased serious society anxiety and led to a higher mortality rate. In order to accurately quantify deaths, our modal considered percentage of elder people in the UK at a ratio *O*, occupancy of available NHS hospital beds over time at a ratios H_t_ and their availability for COVID-19 critical cases at a ratio J_t_. We assumed that critical cases at non-hospital places led to death in 4 days; elderly people in critical condition at hospital led to death in 14 days, and non-elderly people in critical condition at hospital led to death in 21 days.^19^

One parameter was defined to measure intervention intensity over time as M_t_. which was presented by average number of contacts per person per day. We assumed that transmission ratio *β* equals to the product of intervention intensity M_t_ and the probability of transmission (b) when exposed (i.e., *β*= mb). In Wuhan, intervention intensity was assumed within [3-15], and gave with a relatively accurate estimation of COVID-19 breakouts.^13^ We calibrated its value with respect to the population density and human mobility in London and the UK, and estimated outcomes of COVID-2019 outbreaks by implementing different interventions.

All data and code required to reproduce the analysis is available online at: https://github.com/TurtleZZH/Comparison-of-Multiple-Interventions-for-Controlling-COVID-19-Outbreaks-in-London-and-the-UK

### Data sources and modal calibration

Considering that COVID-19 breakouts in Wuhan nearly ended by taking suppression intervention, our model was first fitted and calibrated with data on cases of COVID-19 in Wuhan.^13^ In Fig.1 it showed how suppression (M = 3) impacted on the total number of infections and deaths over time during January 2020 and April 2020. In comparing to other strategies, it demonstrated that the total infections of Wuhan greatly reduced and led an earlier peak time on the 42^nd^ day (Feb 2^nd^ 2020. The end time of releasing suppression was due to the 123^th^ day (April 23^rd^ 2020). It showed that mitigation (M=6) in Wuhan on the 32^nd^ day may lead to 5-6 times more total infections than suppression, although it would delay the outbreak. If Wuhan took a 2 weeks rolling mitigation and suppression intervention (M = 6 or 3), the total infections might be increased 1.5 – 2 times, but the peak time would be delayed and released healthcare demand accordingly.

Using Wuhan’s data, our estimation was close to the practical trend of outbreaks in Wuhan, and gave similar results to other works.^13.22^ We tested that transmission rate from I to S is about 0.157; transmission rate from E to S is about 0.787.^13^ The incubation period was assumed to be 6 days.^8^ As for other parameters, we followed the COVID-19 official report from WHO^19^, and gave a medium estimation on average durations related from infectious, to mild or critical case, and death or recovery were shown in Table.1.

**Table 1:** Parameters estimation in our model.

| Name | Representation | Value | Ref |
| --- | --- | --- | --- |
| **N** | UK population by Aug 2019 | 6.6 million | [20] |
| **i** | Efficiency of isolation contacts | 0.88-1.00 | Tested |
| **$\beta_1$** | Transmission rate from I to S | 0.157 | [13] |
| **$\beta_2$** | Transmission rate from E to S | 0.787 | [13] |
| **$\alpha_1$** | Incubation period | 6 days | [8] |
| **$\alpha_2$** | Average period from M to C | 7 days | [19] |
| **$\gamma_1$** | Average period from E to R | 3.5 days | Assumed |
| **$\gamma_2$** | Average period from M to R | 7 days | [19] |
| **$\gamma_3$** | Average period from Non-H to R | 42 days | Assumed |
| **$\gamma_4$** | Average period of older people from H to R | 18 days | Assumed |
| **$\gamma_5$** | Average period from non-older people from H to R | 13 days | Assumed |
| **$d_1$** | Average period from Non-H to D | 4 days | Assumed |
| **$d_2$** | Average period of older people from H to D | 14 days | [19] |
| **$d_3$** | Average period of non-older people from H to D | 21 days | [19] |
| **m** | Proportion of Mild case | 0.80 | [19] |
| **s** | Proportion of Severe case | 0.138 | [19] |
| **c** | Proportion of Critical case | 0.061 | [19] |
| **$B_t$** | Number of hospital beds in the UK | 167589 | [22] |
| **O** | Percentage of people over 65 in the UK | 0.18 | [21] |
| **$H_t$** | Percentage of unoccupied hospital beds | 0.20-0.60 | Assumed |
| **$J_t$** | Percentage of available hospital beds for COVID-19 critical cases | 0.8-1 | Assumed |
| **$M_t$** | The intensity of intervention | 3-15 | [13] |

Regard as the percentage of elderly people in the UK, it was assumed as 18%.^21^ The total number of NHS hospital beds was given as 167589 with an initial occupied ratio up to 85%.^22^ Considering that UK government began to release NHS hospital beds after COVID-19 breakouts, we assumed the occupied ratio reduced to 80% and would further fall to 40% by April 04, 2020. Accounting for other serious disease cases requiring NHS hospital beds in the early breakout of COVID-19, we assumed that a ratio of available hospital beds for COVID-19 critical cases was initially at 80%, and gradually raised to 100%.

The intervention intensity was related to the population density and human mobility. We gave an initialization to London and non-London regions: London (M=15, population: 9.3 million), non-London regions (M=15, population: 57.2 million). After taking any kind of interventions, we assumed the change of M would follow a reasonable decline or increase in 3-5 days.

### Procedure

Due to difference of population density between London and other regions in the UK, we observed a fact that the accumulative infections in London was about one third of the total infectious population in the UK.^22^ We separately combined the calibrated model with data on the cases of COVID-19 in London, the UK (non-London) and the UK during February 2020 and March 2020 to estimate the total number of infections and deaths, and also peak time and value of healthcare demand by applying different interventions. In contain stage, we assumed a strategy of isolation contacts were taken in the UK from 6^th^ Feb 2020 to 12^th^ March 2020, the effectiveness of isolation of cases and contacts was assumed as 94% in London and 88% in non-London regions.

The key tuning operation was to adjust intensity level of M_t_ over time. We assumed that suppression intensity was given to reduce unaltered internal mobility of a region, where: M = 3. Mitigation intensity was given a wide given range [4-12], where high intensity (M = 4 or 5), moderate intensity (M = 6-8), low intensity (M = 9-12), We evaluated effectiveness of multiple interventions in London and non-London regions, including: suppression, mitigation and rolling intervention. The evaluation metric included 9 indicators as follow: 1: Unimodal or multimodal distribution. 2 If outbreak ends in one year. 3. Total infections. 4. Total deaths. 5. Peak time of healthcare demand. 6. Peak value of healthcare demand for severe and critical cases. 7. Peak time of non-hospital population. 8. Peak value of non-hospital population. 9. Final morality rate (equals to Total deaths over Total infections). The length of intervention was calculated due the date that daily new infections were nearly clear.

Respect to definition of optimal interventions, we first conducted a condition that COVID-19 outbreaks ended as early as possible, and definitely not lasted over 1 year, otherwise it consistently impacted on economic recovery. The second condition was a good balance between total infections/deaths and intervention intensity. The last one was later peak time and smaller peak value of healthcare demand, where it gave sufficient time to prepare essential health sources.

## Results

### Effectiveness of suppression

We estimated that suppression with intensity M = 3 was taken in both London and non-London regions in the UK on the 46^th^ day (March 23^rd^, 2020). The model reproduced the observed temporal trend of cases within London, non-London and the UK. As shown in Fig.3, it captured the exponential growth in infections between the 35^th^ day (March 12^th^, 2020) and the 55^th^ day (April 1^st^, 2020). We estimated that at the day (on March 23^th^, 2020) to take intervention, daily infectious population (Exposed) in the UK actually reached 78579. Our results suggested there were nearly 11 times more infections in the UK than were reported as confirmed cases (6650 on March 23^rd^, 2020).

**Figure 3:**
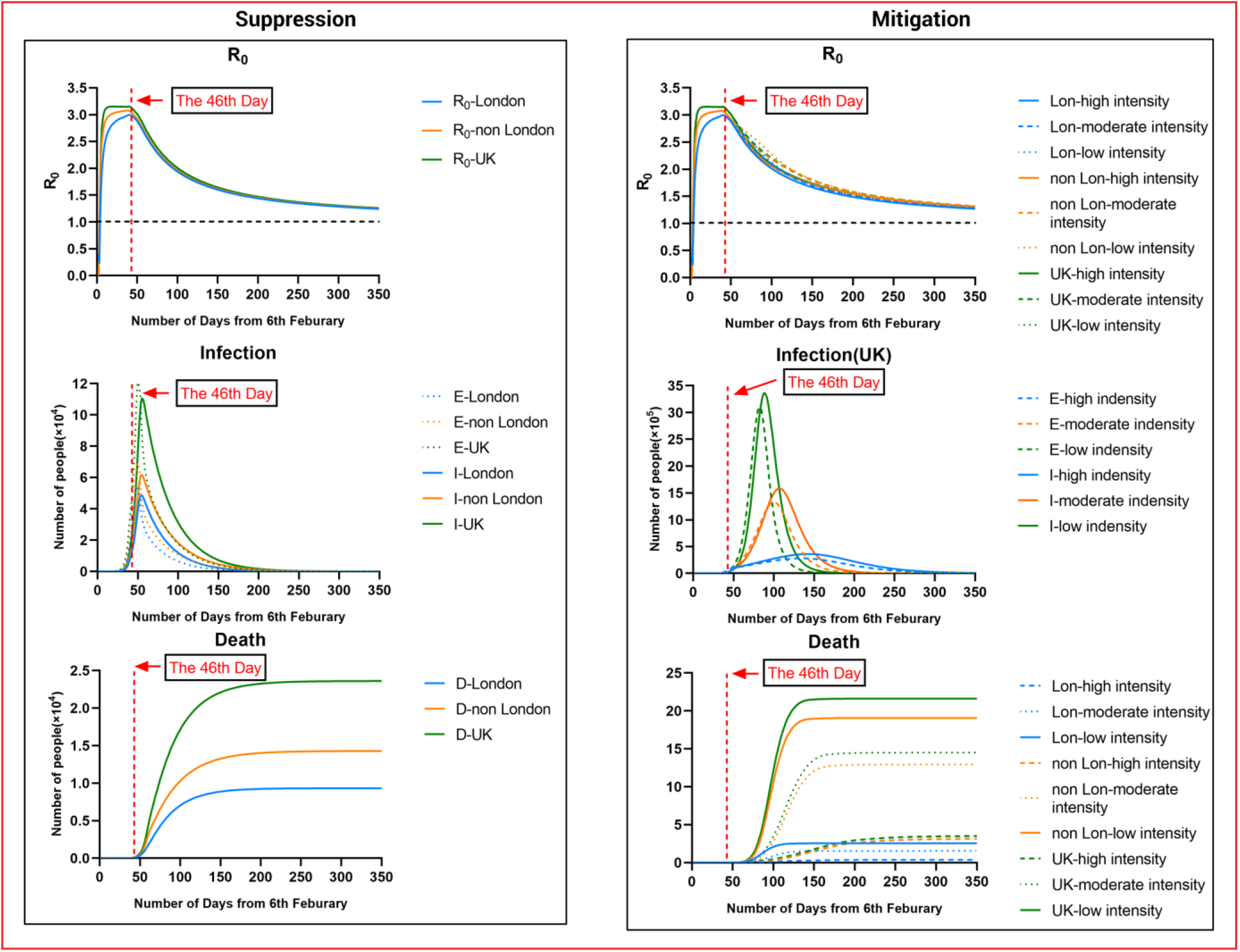
Illustration of controlling COVID-19 outbreaks in London and non-London regions by taking suppression and mitigation with parameters. (a) London population: 9.30 million; non-London population: 57.2 million. (b) Suppression Intervention (M = 3), Mitigation Intervention: Low (M = 10). Moderate (M = 8). High (M =6). (c) Effectiveness of isolation in contact phase (before 12^th^ March 2020): London. 94%, non-London: 88%.

The infections in London nearly occupied about 51% of the overall UK infections. After implementing suppression, the results in the UK appeared a similar trend as Wuhan in Fig.1, where daily exposed and infectious population were greatly reduced. The total deaths by the 200^th^ day (August 24^th^, 2020) in the UK was about 23805, where London had about 9388 deaths and non-London regions had about 14117 deaths. The outbreak of COVID-19 could be possibly controlled by the 100^th^ day (May 16^th^ 2020), and can be nearly ended by the 150^th^ day (July 5^th^ 2020). The difference was that the peak of daily infectious population (E = 54760) of London was nearly 3.4 times greater than the one in Wuhan (E = 15870); the peak time (the 50^th^ day) of daily infections in London was 14 days later than the one (the 36^th^ day) in Wuhan. It was probably because suppression applied in Wuhan (the 32^nd^ day) was 3 days earlier than London (the 35^th^ day). It implied that earlier suppression could reduce infections significantly, but may lead to an earlier peak time of healthcare demand.

We estimated that the predicted R_t_ of London, non-London and the UK dramatically raised in the first 7 days to 2.5 above, and varied from the 2^nd^ days (February 8^th^ 2020) to from the 46^th^ day (March 23^th^ 2020), with values ranging from 2.5 to 3.2. Notably, non-London regions had slightly higher value of R than London during these days. That was because the total population in non-London regions was about 5 times more than the figure in London, as a result of more susceptible and exposed population in non-London regions. After taking suppression in the UK, we estimated a rapid decline in R in later March, from 3 at the 46^th^ day (March 23^th^ 2020) to 1.4 at the 230^th^ day (September 23^rd^ 2020).

### Effectiveness of mitigation

We simulated that mitigation with low, moderate and high intensity (M = 6, 8, 10) were taken in both London and non-London regions in the UK at the 46^th^ day (March 23^rd^, 2020), as show in Fig.3. Considering that the UK went to delay phase on the 35^th^ day (March 12^th^, 2020), M in the UK was adjusted to 12 from March 12^th^ 2020 to March 23^th^ 2020.

The results showed that mitigation strategies were able to delay the peak of COVID-19 breakouts but ineffective to reduce daily infectious populations. We estimated that the peak of daily infectious population was reduced to 3.10 million (M = 10) to 1.33 million (M = 8) or 0.28 million (M = 6); the peak date of daily infections was about on the 82^th^, 100^th^ and 135^th^ day. Compared to suppression, the total deaths in the UK increased to 2.17 million (M = 10) to 1.47 million (M = 8) or 37 thousand (M = 6), where London had about 0.27 million (M = 10) to 165 thousand (M = 8) or 41 thousand (M = 6) and non-London regions had about 1.90 million (M = 10) to 1.30 million (M = 8) or 330 thousand (M = 6). The periods of breakouts with varied mitigations were extended to 180, 200 or 300 days.

Compared to suppression, mitigation taken in the UK gave a slower decline in R in late March, from 3 on the 46^th^ day (March 23^rd^ 2020) to 1.4 on the 280^th^ day (November 12^nd^ 2020). It implied that during this period, there were more infections in the UK. But London had lower R than non-London regions; it implied that London probably would reach a certain level of “herd immunity” earlier.

Above simulations appeared similar trends as findings,^4^ taking mitigation intervention in the UK enabled reducing impacts of an epidemic by flattening the curve, reducing peak incidence and overall death. While total infectious population may increase over a longer period, the final mortality ratio may be minimised at the end. But as similar as taking suppression, mitigation need to remain in place for as much of the epidemic period as possible.

### Effectiveness of multiple interventions

We simulated two possible situations in London and the UK by implementing rolling interventions as shown in Fig. 4. We assumed that all regions in the UK implemented an initial 3 weeks suppression intervention (M=3) from the 46^th^ day (March 23^rd^ 2020) to the 67^th^ day (April 13^rd^ 2020). Then, two possible rolling interventions were given: 1) to keep suppression in London, and take a 3 weeks rolling intervention between suppression and high intensity mitigation (M = 5) in non-London regions; (2) to take a 3 weeks rolling intervention between suppression and high intensity mitigation (M = 5) in all UK.

**Figure 4:**
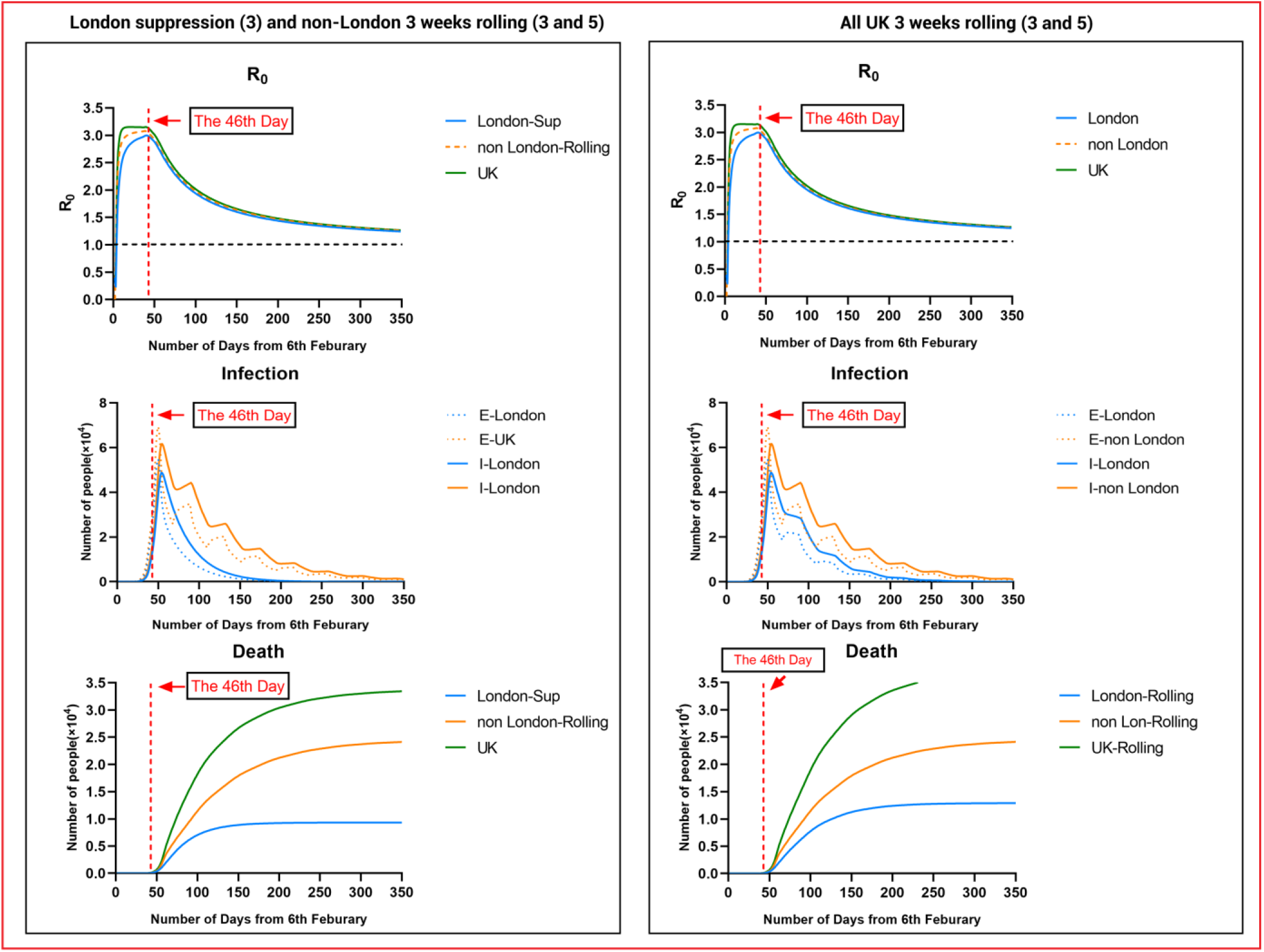
Illustration of controlling COVID-19 outbreaks in London and non-London regions by taking suppression and 3 weeks rolling intervention with parameters. (a) London population: 9.30 million; non-London population: 57.2 million. (b) Suppression Intervention (M = 3), 3 weeks rolling intervention: M = 3-5-3-5, M = 3-4-3-4-3-4. (c) Effectiveness of isolation in contact phase (before 12^th^ March 2020): London. 94%, non-London: 88%.

The simulated results showed the epidemic appeared a unimodal distribution trend over 350 days, longer than the period of suppression. Similar to suppression in Fig.3, the peak date of infectious population in London or non-London regions remain same at the 50^th^ day. After three weeks, rolling intervention with released intensity in non-London regions led to a fluctuation with 4 or 5 peaks of infections until the end of epidemic. The total deaths and infectious population in the UK were greatly reduced to a range from 33 thousand to 37 thousand. It was about 37% - 54% more than the outcome of taking suppression in all the UK.

Above two rolling interventions taken in the UK gave a similar trend of R as suppression, where there was a fast decline in R in late March, from 3 on the 46^th^ day (March 23^rd^ 2020) to 1.4 on the 230^th^ day (September 23^rd^ 2020). It implied that 3 weeks rolling intervention (M = 3 or 5) had equivalent effects on controlling transmissions as suppression, but need to be maintained in a longer period of 350 days.

### Optimal rolling intervention

We simulated other possible rolling interventions with varied period (2, 3 and 4 weeks) and intensity (M = 4, 5 and 6), as shown in Table.2. The results first revealed that rolling intervention with middle intensity (M = 6) cannot control the outbreaks in one year, where the distribution of epidemic was a multimodal trend as similar to mitigation outcomes in Fig.3. The overall infections and deaths significantly increased to over 450 thousand and 60 thousand. While the peak time of healthcare demand for severe critical cases delayed to the 80^th^ – 110^th^ day, the total deaths of the UK would be double than other rolling interventions with low intensity.

**Table 2:** Performance comparison of rolling interventions in the UK. (FMR: Final morality rate = Total deaths / Total infections. PHSC: Peak value of healthcare demand (Severe and Critical cases), PTSC: Peak time of healthcare demand; PnH: Peak value of non-hospital population, PTnH: Peak time of non-hospital population; TD: Total deaths (UK), TI = Total infections (UK), E: End in 1 year, D: Distribution (Unimodal/Multimodal))

| Multiple interventions<br>(intensity 3-5) | U/M | E | TI | TD | PTSC | PHSC | PTnH | PnH | FMR |
| --- | --- | --- | --- | --- | --- | --- | --- | --- | --- |
| **All UK suppression** | **U** | **Y** | **1686778** | **24152** | **64** | **25350** | **57** | **3804** | **1.42%** |
| All UK 3 weeks rolling (3 and 4) | U | Y | 2003228 | 28320 | 64 | 25350 | 57 | 3804 | 1.41% |
| All UK 2 weeks rolling (3 and 5) | U | Y | 2781903 | 38537 | 66 | 25400 | 57 | 3804 | 1.38% |
| **All UK 3 weeks rolling (3 and 5)** | **U** | **Y** | **2699160** | **37432** | **64** | **25350** | **57** | **3804** | **1.38%** |
| All UK 4 weeks rolling (3 and 5) | U | Y | 2612146 | 36288 | 65 | 25360 | 57 | 3804 | 1.38% |
| All UK 2 weeks rolling (3 and 6) | M | N | 4654493 | 62838 | 82 | 29770 | 57 | 3804 | 1.35% |
| All UK 3 weeks rolling (3 and 6) | M | N | 4611192 | 62082 | 96 | 27680 | 57 | 3804 | 1.34% |
| All UK 4 weeks rolling (3 and 6) | M | N | 4501602 | 60670 | 110 | 25780 | 57 | 3804 | 1.34% |
| All UK 3 weeks rolling (3 and 8) | M | N | 16122311 | 257848 | 181 | 96020 | 181 | 27000 | 1.59% |
| London suppression (3), other regions 2 weeks rolling (3 and 5) | U | Y | 2484942 | 34621 | 65 | 25380 | 57 | 3804 | 1.39% |
| **London suppression (3), other regions 3 weeks rolling (3 and 5)** | **U** | **Y** | **2430060** | **33884** | **65** | **25350** | **57** | **3804** | **1.39%** |
| London suppression (3), other regions 4 weeks rolling (3 and 5) | U | Y | 2370997 | 33110 | 65 | 25360 | 57 | 3804 | 1.39% |

Another finding was that given equivalent intensity (M= 3 or 5) of rolling interventions, the longer period (4 weeks) led to slight reduction of the total deaths to 36288, compared to 37432 of 3 weeks rolling and 38537 of 2 weeks rolling in the UK. The peak time of healthcare demand nearly occurred at same: the 64^th^-65^th^ day; with an equivalent peak value. Thus, in balance of total deaths and human mobility restriction, 3 weeks of period might be a feasible choice.

We considered the length of intervention in the UK impacting on social and economic. Maintaining a period of suppression in London, it was possible to control the outbreaks at the 100^th^-150^th^ day that minimized economic loss to the greatest extent. Due to lower population density and less human mobility of non-London regions, 3 weeks rolling intervention was appropriated to non-London regions for balancing the total infections and economic loss, but the length of this strategy was extended to 300 days.

## Discussion

Aiming at a balance of infections, deaths and economic loss, we simulated and evaluated how and when to take which intensity level of interventions was a feasible way to control the COVID-19 outbreak in the UK. We found rolling intervention between suppression and mitigation with high intensity could be an effective and efficient choice to limit the total deaths of the UK to 33-38 thousand but maintain essential mobility for avoiding huge economic lose and society anxiety in a long period. Rolling intervention was more effective to non-London regions in the UK. Due to lower population density and less human mobility, realising some intervention intensity would not lead to a second breakout of COVID-19 and benefit maintenance of business activities. Considering difference and diversity of industrial structure of London and non-London regions in the UK, hybrid intervention was more suitable and effective to control outbreaks. It could complete London outbreak with suppression in 3 months, and tolerate a longer recovery period of non-London regions taking 3 weeks rolling intervention. The rapid completion of outbreak in London would strongly benefit to economic recovery of the UK. Other regions maintained essential production and business activities to offer sufficient support to London during these 3 months. After 3 months, London could drive UK economic recovery and help other regions of the UK.

In above scenarios, our model found that the total infections in the UK was limited to 2.35 million; the total deaths in the UK was limited to 33.4 thousand. Also, the peak time of healthcare demand would occur at the 65^th^ day (April 11^th^ 2020), where it needed sufficient hospital beds to accommodate 25.3 thousand severe and critical cases. This scenario echoed that applying suppression at a right time was crucial to delay the peak date of healthcare needs and increase available hospital beds for severe and critical cases. We found that while immediate suppression being taken in Wuhan at 3 days earlier than London reduced 3.4 times infections, it led to nearly 1.56 times of severe and critical cases at non-hospital places (Wuhan: Peak 1389 at the 43^th^ day, London, Peak 889 at the 57^th^ day). It implied that taking immediate suppression without sufficient hospital beds was risky and led to more deaths in the early breakout.

Our finding revealed that implementing suppression intervention required considering other conditions of this region like culture difference, industrial structure, etc. Success of immediate suppression in Wuhan relied on strict lockdown of human mobility to community level and sufficient resource support from other cities or provinces in China. If there were no sufficiently external support, it would be risky to take highly intensive suppression to entire country due to shortage of healthcare resources and huge impacts on its economics. In the UK, it was hardly to practically implement the same level of intensity as Wuhan. If intensive suppression was relaxed at any time points, the transmission would quickly rebound. This was more like a multi-modal curve when taking multi-intervention strategies in Fig.1. Therefore, we concluded that taking rolling intervention was more suitable to the UK.

Notably, the total infections estimated in our model was measured by Exposed population (asymptomatic), which might be largely greater than other works only estimating Infectious population (symptomatic). We found that a large portion of self-recovered population were asymptomatic or mild symptomatic in the COVID-19 breakouts in Wuhan (occupied about 42%-60% of the total infectious population). These people might think they had been healthy at home because they did not go to hospital for COVID-19 tests. It was one important issue that some SEIR model predicted infectious population in Wuhan that 10 times over than confirmed cases.^12.13^ Early release of intensity might increase a risk of the second breakout.

There are some limitations to our model and analysis. First, our model’s prediction depends on an estimation of intervention intensity that is presented by average-number contacts with susceptible individuals as infectious individuals in a certain region. We assumed that each intervention had equivalent or similar effect on the reproduction number in different regions over time. The practical effectiveness of implementing intervention intensity might be varied with respect to cultures or other issues of certain county. In the UK or similar countries, how to quantify intervention intensity needs an accurate measure of combination of social distancing of the entire population, home isolation of cases and household quarantine of their family members. As for implementing rolling interventions in the UK, the policy needs to be very specific and well-estimated at each day according to the number of confirmed cases, deaths, morality ratio, health resources, etc. Secondly, our model used a variety of plausible biological parameters for COVID-19 based on current evidence as shown in Table.1, but these assumed values might be varied by populations or countries. For instance, we assumed that average period of mild cases to critical cases is 7 days, and average period of elderly people in hospital from severe cases to deaths was 14 days, etc. The change of these variables may impact on our estimation of infections and deaths in the UK. Lastly, our model assumes a condition that there will be a reasonable growth of available hospital source as time goes in the UK after 23^rd^ March 2020. This was actually supported by latest news that Nightingale hospital that enables holding 4000 patients opened at London Excel centre on 4^th^ April 2020.^24^

Our results show that taking rolling intervention is one optimal strategy to effectively and efficiently control COVID-19 outbreaks in the UK. This strategy potentially reduces the overall infections and deaths; delays and reduces peak healthcare demand. In future, our model will be extended to investigate how to optimise the timing and strength of intervention to reduce COVID-19 morality and specific healthcare demand.

## Data Availability

All data and code required to reproduce the analysis are available online.

https://github.com/TurtleZZH/Comparison-of-Multiple-Interventions-for-Controlling-COVID-19-Outbreaks-in-London-and-the-UK

## References

[1] World Health Organization. Coronavirus disease 2019 (COVID-2019): situation report—73. April 2nd, 2020. https://www.who.int/emerge-ncies/diseases/novel-coronavirus-2019/situation-reports (accessed April 3rd, 2020).

[2] Anderson RM, Heesterbeek H, Klinkenberg D, Hollingsworth TD. Comment How will country based mitigation measures influence the course of the COVID-19 epidemic? The Lacnet, 2020;2019(20):1–4

[3] Imperial College London, MRC Centre for Global Infectious Disease Analysis. News COVID-19—report 12: The global impact of COVID- 19 and strategies for mitigation and suppression. Mar 26, 2020 https://www.imperial.ac.uk/mrc-global-infectious-disease-analysis/covid-19/report-12-global-impact-covid-19/. (accessed March 28, 2020).

[4] Imperial College London, MRC Centre for Global Infectious Disease Analysis. News / COVID-19—report 9: Impact of non-pharmaceutical interventions (NPIs) to reduce COVID-19 mortality and healthcare demand. Mar 16, 2020. https://www.imperial.ac.uk/mrc-global-infectious-disease-analysis/covid-19/report-9-impact-of-npis-on-covid-19/ (accessed March 18, 2020).

[5] Tian HY, Liu YH, Li YD, Wu CH, Chen B, Kraemer MUG, Li BY, Cai J, Xu B, Yang QQ, Wang B, Yang P, Cui YJ, Song YM, Zheng P, Wang QY, Bjornstad ON, Yang RF, Grenfell BT, Pybus OG and Dye C. Report An investigation of transmission control measures during the first 50 days of the COVID-19 epidemic in China. Science 2020; March 31: e1001076, DOI: 10.1126/science.abb6105

[6] Goldman Sachs, News: Goldman Sees China’s Economy Slumping 9% in First Quarter, 17th March 2020. https://www.bloomberg.com-/news/articles/2020-03-17/goldman-now-see-china-s-economy-slumping-9-in-first-quarter (accessed March 28th, 2020).

[7] Hellewell J, Abbott S, Gimma A, et al. Feasibility of controlling COVID-19 outbreaks by isolation of cases and contacts. Lancet Glob Health 2020;

[8] Backer JA, Klinkenberg D, Wallinga J. The incubation period of 2019- nCoV infections among travellers from Wuhan, China. medRxiv 2020; published online Jan 28. DOI:10.1101/2020.01.27.20018986 (preprint).

[9] Abbott S, Hellewell J, Munday J, CMMID nCoV working group, Funk S. The transmissibility of novel coronavirus in the early stages of the 2019– 20 outbreak in Wuhan: exploring initial point-source exposure sizes and durations using scenario analysis. Wellcome Open Res 2020; 5: 17.

[10] Imai N, Cori A, Dorigatti I, et al. Report 3: transmissibility of 2019- nCov. 2020. https://www.imperial.ac.uk/media/imperial-college/medicine/sph/ide/gida-fellowships/Imperial-2019-nCoV-transmissibility.pdf (accessed Feb 25, 2020).

[11] Mossong J, Hens N, Jit M, Beutels P, et al. Social contacts and mixing patterns relevant to the spread of infectious diseases, PLoS Med, 2008, Mar, 5(3): e74.

[12] Kucharski AJ, Russell TW, Diamond C, et al. Early dynamics of transmission and control of COVID-19: a mathematical modelling study. Lancet Infect Dis [Internet] 2020;3099(20):2020.01.31.20019901.

[13] Zifeng Yang, et.al., Modified SEIR and AI prediction of the epidemics trend of COVID-19 in China under public health intervention., Journal of Thoracic Disease, 2020.

[14] Rachale Pung, et.al., Investigation of three clusters of COVID-19 in Singapore: implications for surveillance and response measures, Lancet. 2020.

[15] Joel Hellewll, et.al., Feasibility of controlling COVID-19 outbreaks by isolation of cases and contacts.

[16] Wu JT, Leung K, Leung GM. Nowcasting and forecasting the potential domestic and international spread of the 2019- nCoV outbreak originating in Wuhan, China: a modelling study. Lancet 2020.

[17] Yang S, et.al., Early estimation of the case fatality rate of COVID-19 in mainland China: a data-driven analysis. Ann Transl Med. 2020 Feb; 8(4):128. doi: 10.21037/atm.2020.02.66.

[18] Abdul Kuddus, Azizur Rahman, MR Talukder, and Ashabul Hoque. 2014. A modified SIR model to study on physical behaviour among smallpox infective populations in Bangladesh. American Journal of Mathematics and Statistics 4, 5 (2014), 231–239

[19] World Health Organization. Report of the WHO-China Joint Mission on Coronavirus Disease 2019 (COVID-19), pp.14. April 2nd, 2020 https://www.who.int/docs/default-source/coronaviruse/who-china-joint-mission-on-covid-19-final-report.pdf. (accessed March 8th, 2020).

[20] Office for National Statistics. News/ Overview of the UK population: August 2019. https://www.ons.gov.uk/peoplepopulationandcomm-unity/populationandmigration/populationestimates/articles/overvie-woftheukpopulation/august2019/previous/v1 (accessed March 8th, 2020).

[21] Office for National Statistics. News/ Living longer: how our population is changing and why it matters. https://www.ons.gov.uk/-peoplepopulationandcommunity/birthsdeathsandmarriages/ageing/articles/livinglongerhowourpopulationischangingandwhyitmatters/2018-08-13 (accessed March 8th, 2020).

[22] Statista Com. Report/ Annual number of hospital beds in the UK from 2000 to 2017. Jan, 2020. https://www.statista.com/statistics-/473264/number-of-hospital-beds-in-the-united-kingdom-uk/ (accessed March 8th, 2020).

[23] Nuffield Trust, Reports/ Hospital bed occupancy, April 26th 2019 https://www.nuffieldtrust.org.uk/resource/hospital-bed-occupancy. (accessed March 8th, 2020).

[24] NHS Professional, News/ NHS Nightingale-London, April 3rd 2020. https://www.nhsprofessionals.nhs.uk/en/Nightingale/Nightingale/Nightingale-London (accessed April 5th, 2020).

